# Incidence and Determinants of Post-Pandemic Catastrophic Health Expenditure Among Households in a Southern State of Malaysia

**DOI:** 10.1101/2025.08.01.25332638

**Authors:** Siti Zulaikha Mohd Zainudin, Sharifah Najwa Syed Mohamad, Nizam Baharom, Mohamed Fakhri Abu Baharin

## Abstract

The COVID-19 pandemic caused an unprecedented burden on healthcare and significantly disrupted household economies that reshaped households’ expenditure patterns, particularly in health-related expenditure. High out-of-pocket health expenditure (OOPHE) inherently exposed households at risk of incurring catastrophic health expenditure (CHE). This study aimed to investigate the incidence and key determinants of CHE in the post-pandemic period among households in a southern state of Malaysia. A cross-sectional survey was conducted in Negeri Sembilan, a southern state of Malaysia between February and August 2024. A total of 400 head of households participated via proportionate stratified sampling. CHE was defined as the amount of OOPHE exceeding 10% of the monthly household’s expenditure. Descriptive analysis was performed to examine the sociodemographic characteristics of household heads and to determine the median monthly OOPHE (in MYR), while regression analysis was conducted to identify the determinants of CHE. The incidence of CHE was 16.8%. The determinants of CHE included smallest household size (aOR;2.874, 95% CI;1.317,6.271), household in Rembau district (aOR;13.191, 95% CI;3.061,56.843), household with higher education (aOR;2.995, 95% CI:1.599,5.609), and those who are not working (aOR;2.566, 95% CI;1.349,4.880). The post-pandemic incidence of CHE among households in Negeri Sembilan, Malaysia was high. Lower household size, district’s location, higher educational level and lack of occupation were significant determinants of CHE. Findings from this study highlight the urgent need for regular monitoring and targeted financial protection strategies to reduce CHE and promote equitable healthcare access in the post-pandemic period.

## Introduction

The COVID-19 pandemic has significantly affected the global healthcare system, and plunged countries into economic crisis, resulting in unparalleled financial burdens (1). Global lockdowns and shutdowns have been implemented to stop the virus from spreading, shutting down many businesses, interrupting global supply chains and putting the foot on the brake pedal for economic activity. In turn, the world economy was placed under severe stress, resulting in workforce reduction and surging unemployment rate. By 2020, the global unemployment rate had increased to 6.5%, which is also a 1.1% increase from the previous year and led to substantial income reduction for households (2). This economic instability caused immense financial strain among households to afford for their daily needs, and led to apparent shift in livelihood, particularly for healthcare.

The pandemic not only intensified healthcare demand but also significantly boosted the healthcare expenditure globally, notably in out-of-pocket health expenditure (OOPHE). Global OOPHE demonstrated a slight increased from 16.33% in 2020 to 17.23% in 2022 (3). Even Malaysia is not isolated from such trend, as the country has demonstrated a rising pattern in OOPHE over the last decade. According to previous report, OOPHE in Malaysia had increased from 31.89 in 2011 to 31.89%, and further escalated to 37.22% by 2022 (4). Comparatively, Malaysia recorded the highest OOPHE (37%) among upper-middle-income countries in Southeast Asia, surpassing Indonesia and Thailand at 32% and 9% respectively (3). High amount of OOPHE carries profound implications, exerting a negative impact on household financial risk protection leading to impoverishment and most importantly, inducing catastrophic health expenditure (CHE).

The definition of CHE refers to the condition where households incur OOPHE that exceed affordability thresholds, commonly defined as spending more than 10% of household income or consumption, or over 40% of capacity to pay after meeting basic needs (5,6). The incidence of CHE varied across countries due to differences between health financing system, economic status, and health policy. Prior to the COVID-19 pandemic, countries with strong financial protection or comprehensive national health insurance often recorded a lower incidence of CHE (7,8). In contrast, countries such as India and Uganda experienced significantly higher incidence of CHE due to the pandemic, which recorded at 49.7% and 37.7% respectively. This was primarily due to limited financial protection mechanisms, low government investment in health and a high dependence on OOPHE, whereas high-income countries generally had lower incidence of CHE as result of better financing for health (9,10).

CHE in Malaysia during pre-pandemic recorded lower incidence at 2.8% in 2016 (11), however during the pandemic the incidence of CHE rose up to 20.2% in 2021 (11,12). CHE incidence is generally greater among high-risk population such as those with chronic diseases. For instance, CHE incurred among 87% of tuberculosis, 54.4% of cancer and 50% of haemodialysis patients (13–15).

The incidence of CHE poses a major threat to household financial security. Households grappled with CHE usually are forced to sacrifice essential needs, incur debt, or fall into poverty to be able to afford healthcare (16,17). This creates a further barrier to healthcare, especially for those who cannot afford to pay directly for medical services. While previous literature on CHE has been conducted during pre-pandemic and pandemic phase, studies specifically addressing CHE in the post-pandemic and its determinants remain notably limited. Therefore, this study aims to investigate the incidence and key determinants of CHE in the post-pandemic among households in a southern state of Malaysia.

## Materials and methods

### Study design and sample population

This cross-sectional study was conducted among households Negeri Sembilan, which located in south of Peninsular Malaysia. This state has a total of seven districts. A proportionate stratified sampling method was applied to ensure representativeness across all districts. According to data from the Department of Statistics Malaysia (DOSM), Negeri Sembilan had approximately 1.24 million in 2023 (18). The sample size needed for this study was 400 respondents. The calculated proportionate samples from each district were: Seremban (211), Port Dickson (44), Jempol (45), Tampin (33), Kuala Pilah (25), Jelebu (20), and Rembau (19). Samples at each stratum were then collected via convenience sampling from February to August 2024.

The inclusion criteria for head of households included were aged 18 years or older, Malaysian nationality, able to understand, read, and write in either Malay or English. The exclusion criteria were households that live other than designated permanent living quarters, such as old folks home, and temporary shelter.

### Study tools

The questionnaire used in this study was adapted from the National Health and Morbidity Survey (NHMS) 2019 (19). This questionnaire comprises three sections: (a) sociodemographic profile, (b) monthly household expenditure, and (c) details of monthly OOPHE. Sociodemographic profile consisted of age, gender, ethnic group, marital status, education level, employment status, household income (monthly, in MYR), household size, number of elderly (60 years old and older), and number of family members with disability.

Monthly households’ expenditure included detail expenditure (in MYR) on 12 groups of expenditure items such as food, utilities, educational fees, health insurance, transportation, communication, health care expenditure and others. Details of monthly OOPHE comprised of private health insurance status, OOPHE details made for pharmaceuticals product, medical appliances and equipment, outpatient-based care and hospital-based care.

The questionnaire was piloted among 50 respondents, with no changes made for the final version. Data collection was conducted via face-to-face interviews.

### Data Analysis

Data was analyzed using IBM Statistical Package for the Social Sciences (SPSS) version 24.0. Descriptive statistics were used to summarize the sociodemographic characteristics of households. OOPHE (MYR) was described in median (IQR). CHE was identified when a monthly household OOPHE exceeded 10% of the total household’s expenditure (3).

Logistic regression analysis was conducted to identify the determinants of CHE. The dependent variable was CHE status, while the independent variable consists of age, household size, gender, ethnicity, district, residence, marital status, educational level, occupation, household income classification, presence of elderly members, household with chronic diseases, households with OKU members and private health insurance status. Backward elimination methods were employed for variable selection. Multicollinearity and interaction were evaluated using correlation matrix, standard error and variance inflation factor including testing for all two-way interactions. Model fit was examined using the Hosmer-Lemeshow test, classification table and ROC curve. Adjusted odds ratios (OR) along with 95% confidence intervals, and *p*-values were reported in the multivariate logistic regression model.

### Ethical Considerations

The ethic approval was obtained from the Human Research and Ethics Committee, Universiti Sains Islam Malaysia. Informed written consent was obtained from all participating respondents. Participants were reassured of the anonymity and confidentiality of personal data.

## Result

A total of 400 respondents from seven districts in Negeri Sembilan participated in the survey. Table 1 shows the sociodemographic characteristics of respondents as well as the corresponding monthly OOPHE (MYR).

**Table 1.**
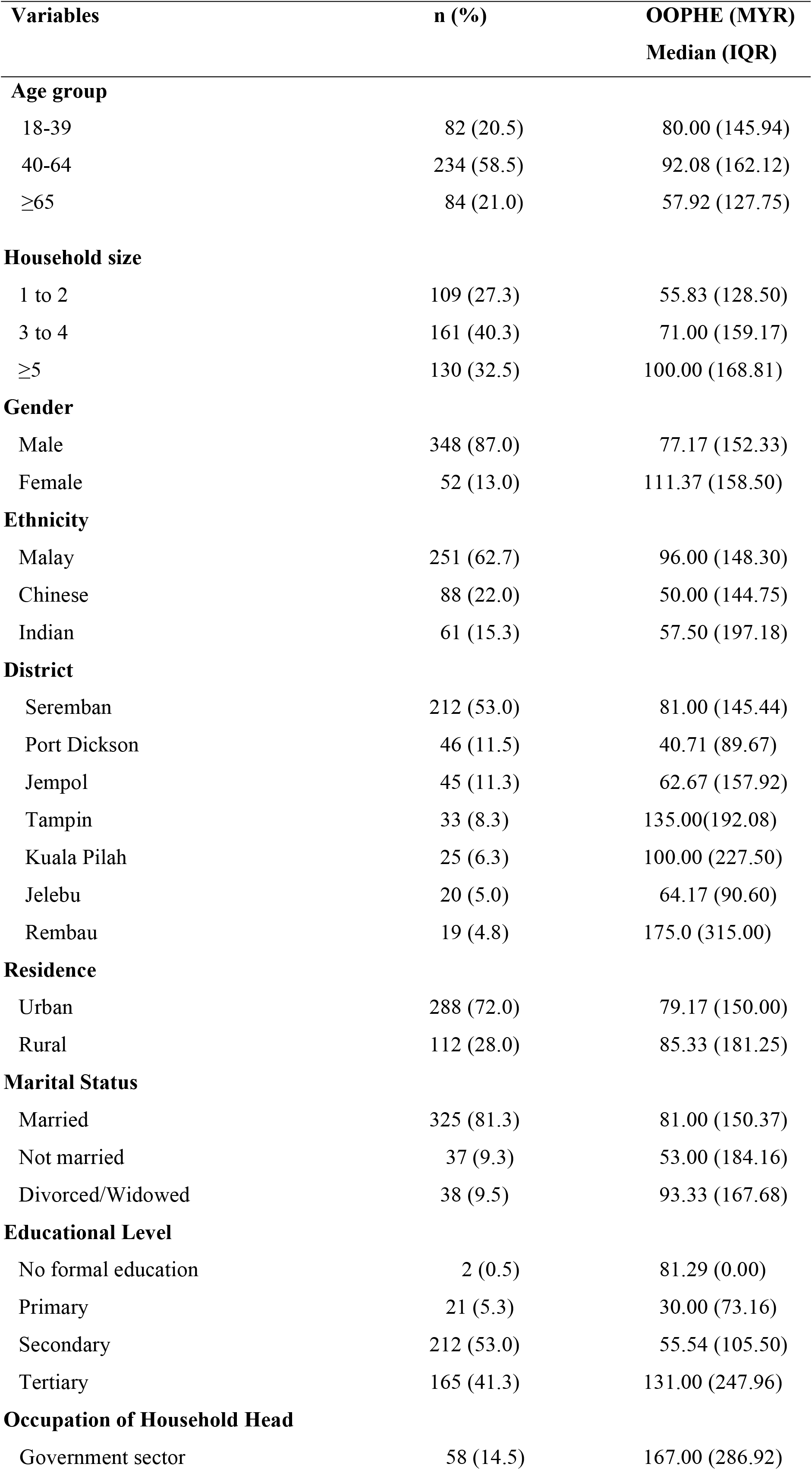
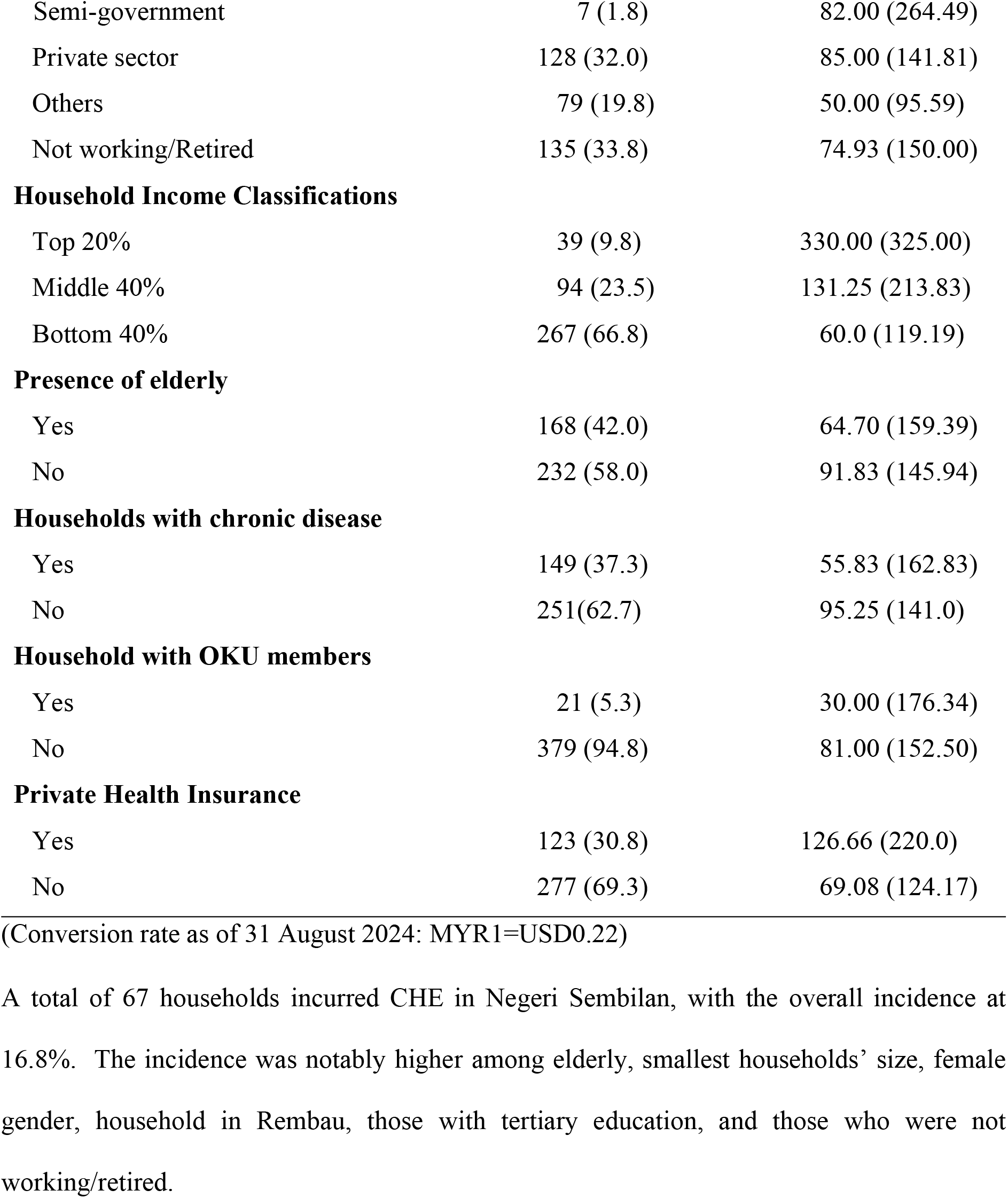
Sociodemographic characteristics and the corresponding monthly household out-of-pocket health expenditure (OOPHE) in Negeri Sembilan (n=400).

**Table 2.**
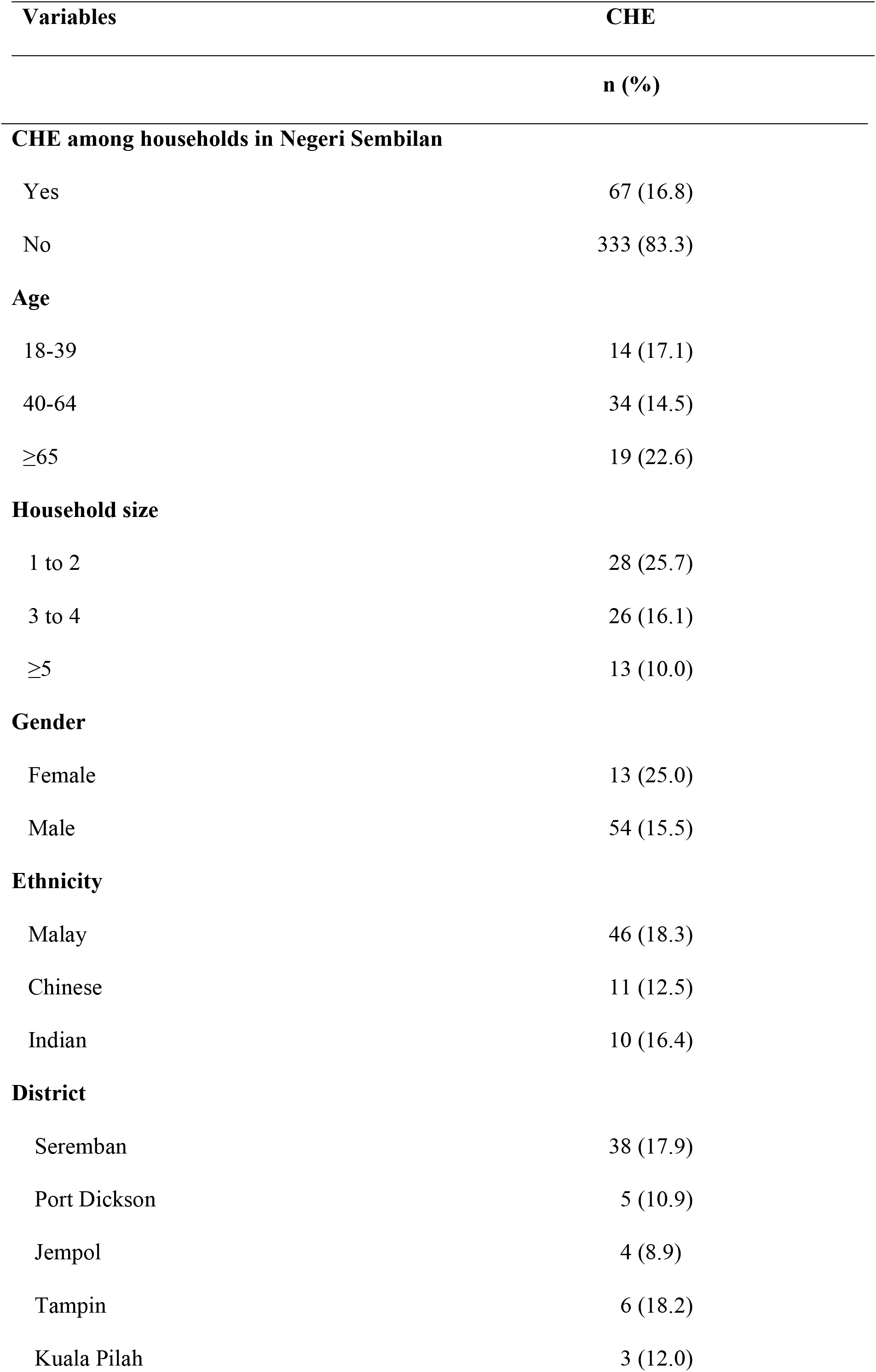
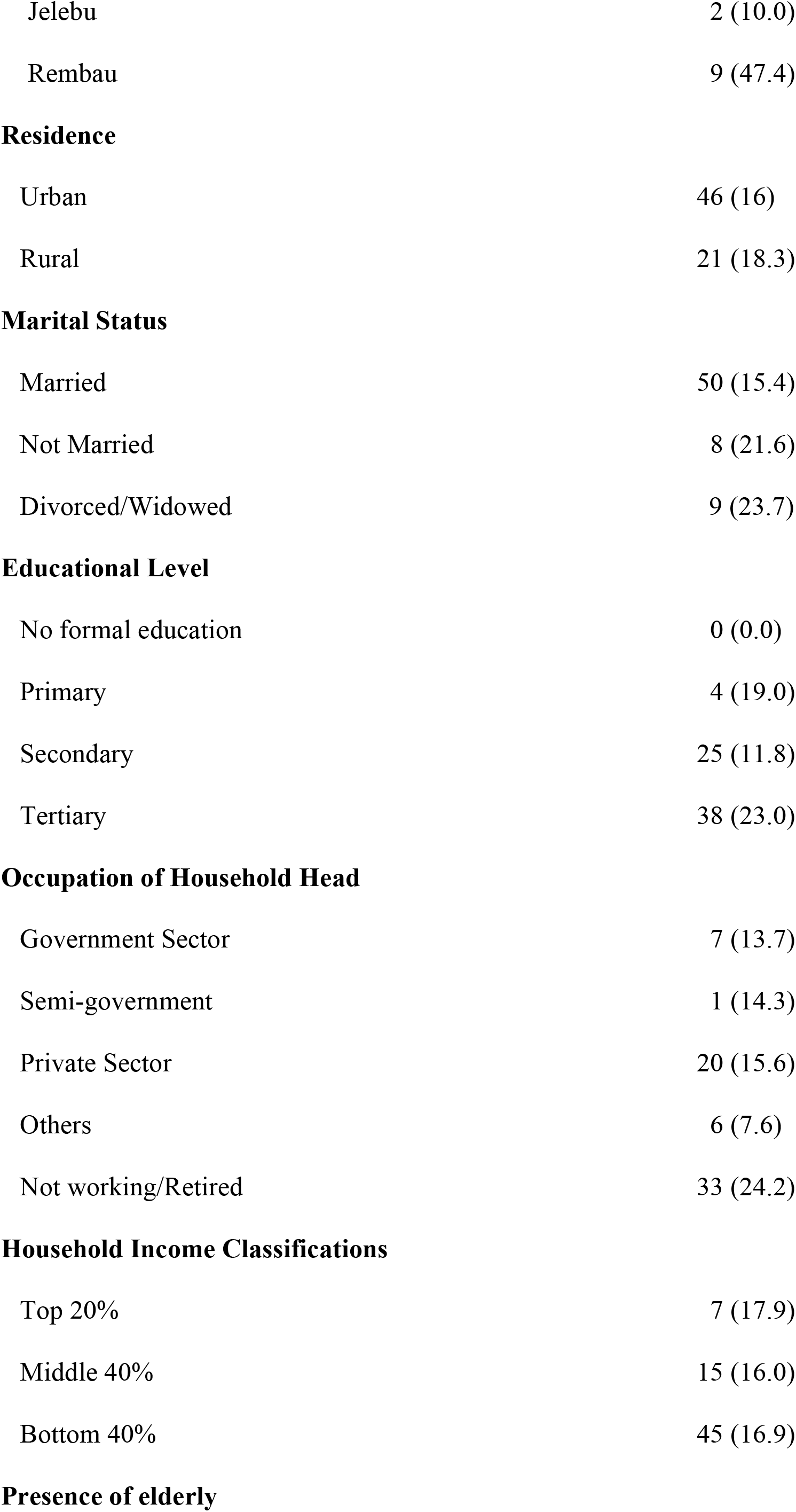
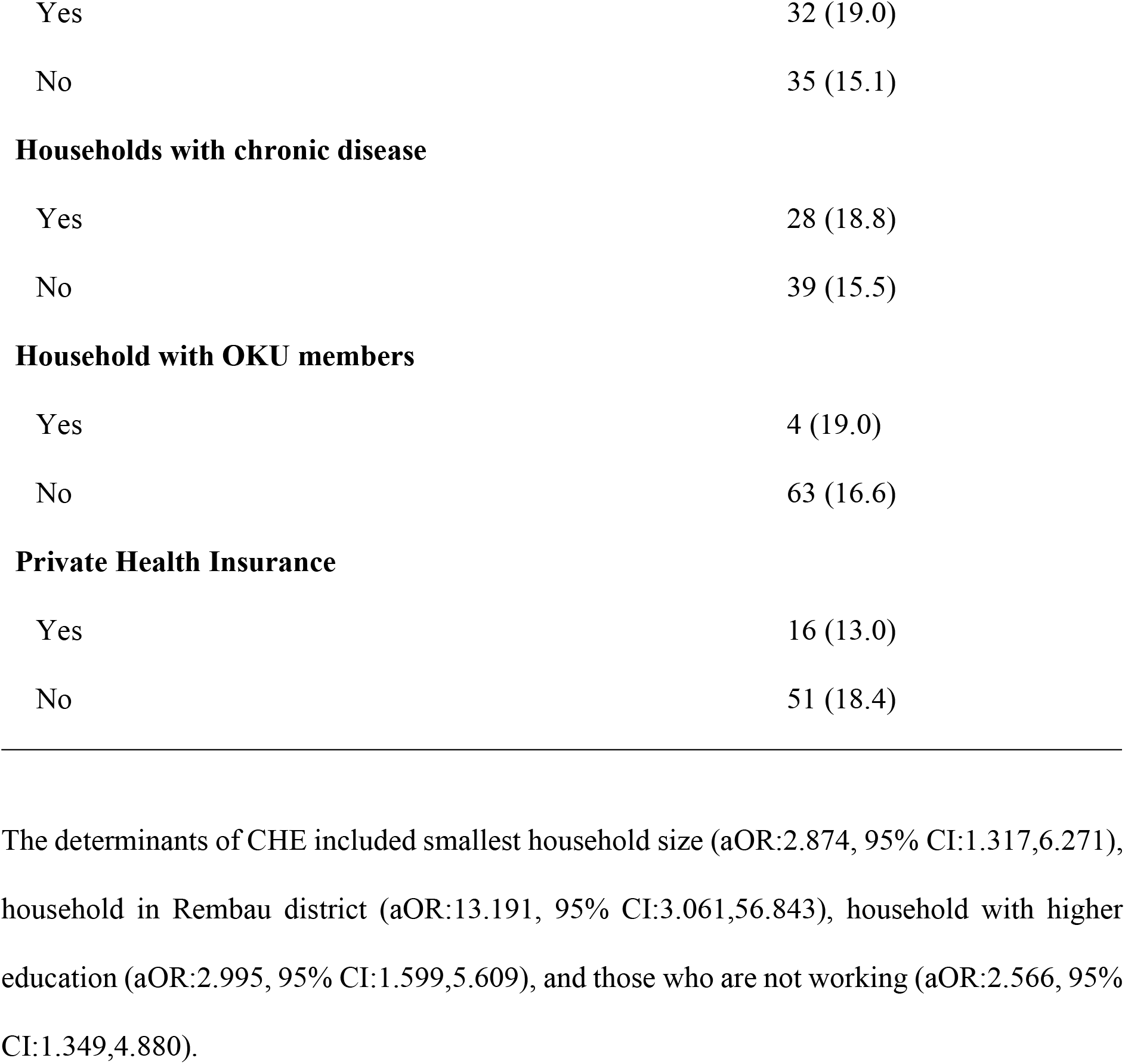
Incidence of catastrophic health expenditure (CHE) based on 10% thresholds according to sociodemographic characteristics of all respondents in Negeri Sembilan (n=67).

| Variables | CHE |
| --- | --- |

|  | n (%) |
| --- | --- |
| **CHE among households in Negeri Sembilan** |  |
| Yes | 67 (16.8) |
| No | 333 (83.3) |
| **Age** |  |
| 18-39 | 14 (17.1) |
| 40-64 | 34 (14.5) |
| ≥65 | 19 (22.6) |
| **Household size** |  |
| 1 to 2 | 28 (25.7) |
| 3 to 4 | 26 (16.1) |
| ≥5 | 13 (10.0) |
| **Gender** |  |
| Female | 13 (25.0) |
| Male | 54 (15.5) |
| **Ethnicity** |  |
| Malay | 46 (18.3) |
| Chinese | 11 (12.5) |
| Indian | 10 (16.4) |
| **District** |  |
| Seremban | 38 (17.9) |
| Port Dickson | 5 (10.9) |
| Jempol | 4 (8.9) |
| Tampin | 6 (18.2) |
| Kuala Pilah | 3 (12.0) |
| Jelebu | 2 (10.0) |
| Rembau | 9 (47.4) |
| **Residence** |  |
| Urban | 46 (16) |
| Rural | 21 (18.3) |
| **Marital Status** |  |
| Married | 50 (15.4) |
| Not Married | 8 (21.6) |
| Divorced/Widowed | 9 (23.7) |
| **Educational Level** |  |
| No formal education | 0 (0.0) |
| Primary | 4 (19.0) |
| Secondary | 25 (11.8) |
| Tertiary | 38 (23.0) |
| **Occupation of Household Head** |  |
| Government Sector | 7 (13.7) |
| Semi-government | 1 (14.3) |
| Private Sector | 20 (15.6) |
| Others | 6 (7.6) |
| Not working/Retired | 33 (24.2) |
| **Household Income Classifications** |  |
| Top 20% | 7 (17.9) |
| Middle 40% | 15 (16.0) |
| Bottom 40% | 45 (16.9) |
| **Presence of elderly** |  |
| Yes | 32 (19.0) |
| No | 35 (15.1) |
| Households with chronic disease |  |
| Yes | 28 (18.8) |
| No | 39 (15.5) |
| Household with OKU members |  |
| Yes | 4 (19.0) |
| No | 63 (16.6) |
| Private Health Insurance |  |
| Yes | 16 (13.0) |
| No | 51 (18.4) |

**Table 3.**
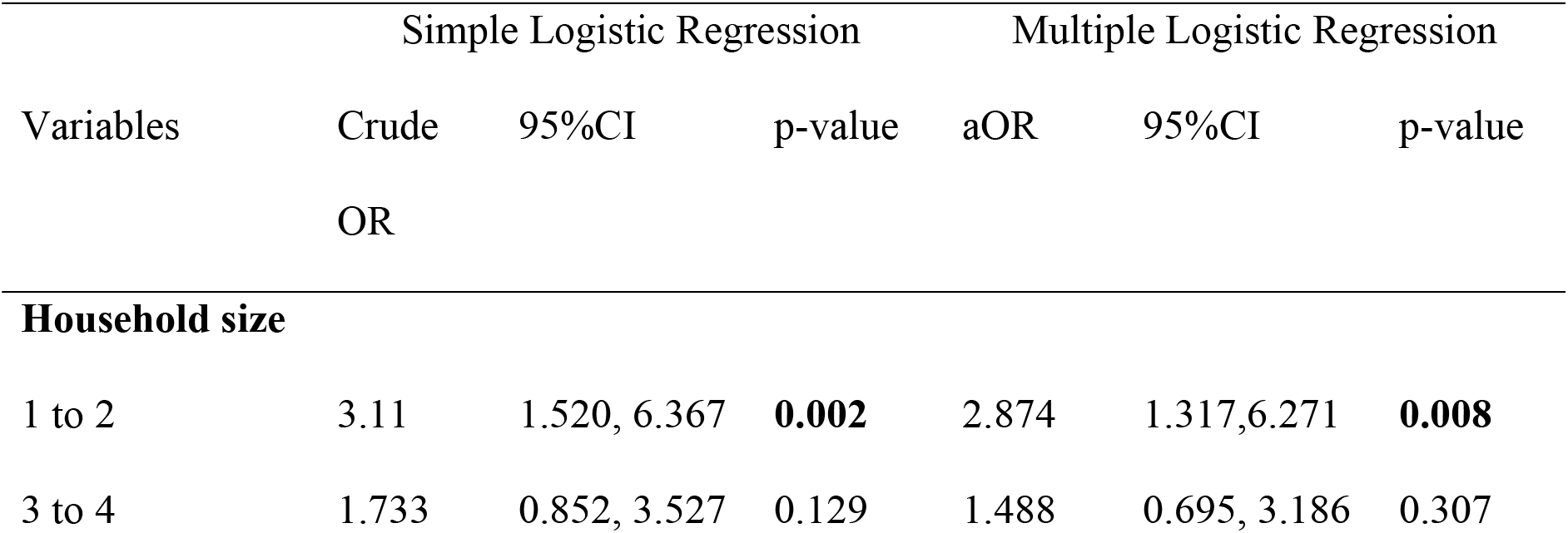
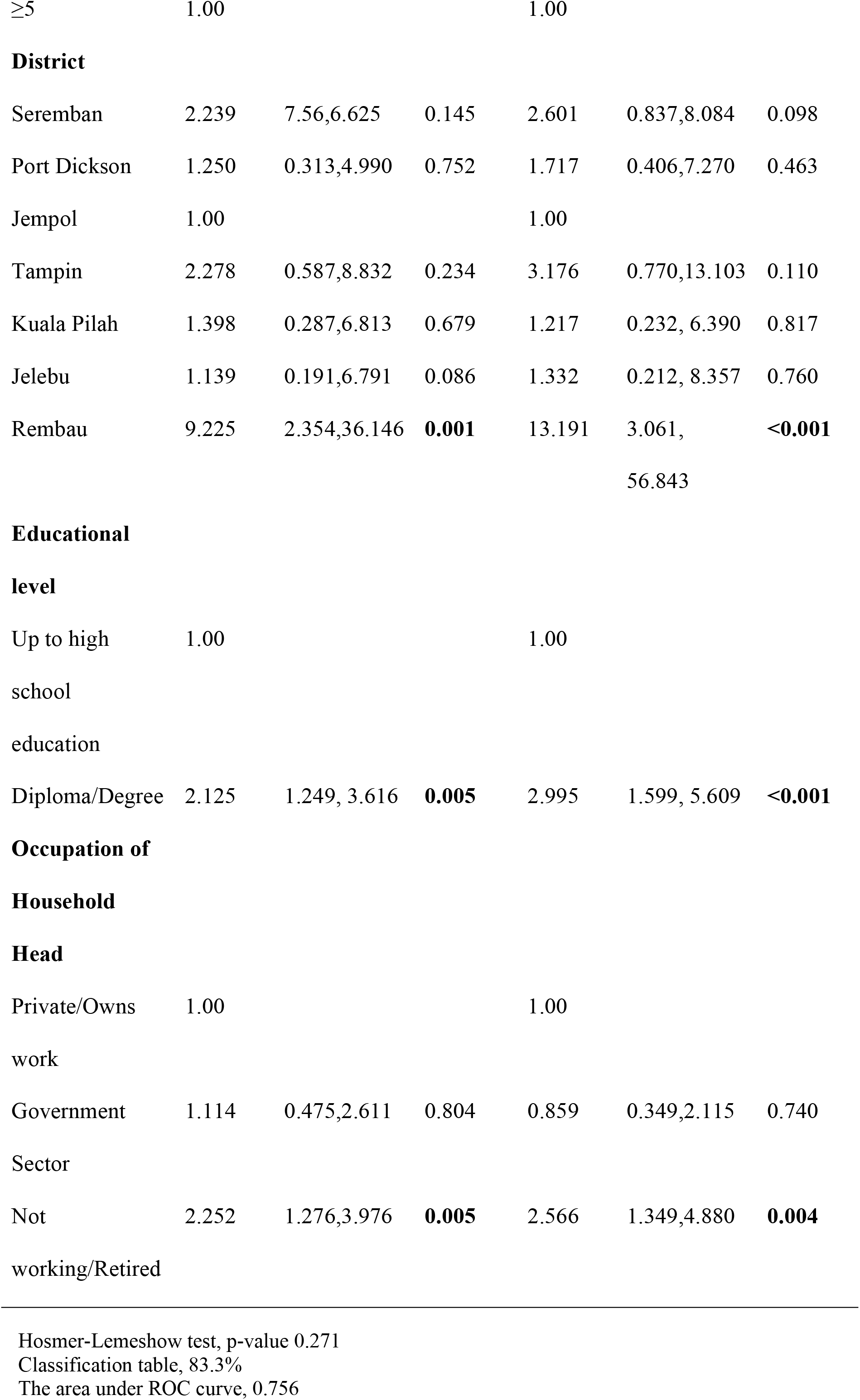
Logistic regressions analysis on determinants of CHE among households in Negeri Sembilan (n=400).

The mean (SD) for age was 52.2 (14.5). Majority of respondents were aged between 40 to 64 years old (58.5%), males (87.0%), of Malay ethnicity (62.7%), resided in urban areas (72.0%), married (81.3%), and household income status from the bottom 40% (66.8%).

The median (IQR) OOPHE for households in Negeri Sembilan were MYR80.50 (152.33). The minimum OOPHE in Negeri Sembilan was MYR1, while the maximum OOPHE was MYR4637.08.

The monthly OOPHE were notably higher among larger household size, female household head, Malay ethnicity, households in the Rembau district, those with tertiary education, those in government sector, high income earners, and those with private health insurance.

A total of 67 households incurred CHE in Negeri Sembilan, with the overall incidence at 16.8%. The incidence was notably higher among elderly, smallest households’ size, female gender, household in Rembau, those with tertiary education, and those who were not working/retired.

The determinants of CHE included smallest household size (aOR:2.874, 95% CI:1.317,6.271), household in Rembau district (aOR:13.191, 95% CI:3.061,56.843), household with higher education (aOR:2.995, 95% CI:1.599,5.609), and those who are not working (aOR:2.566, 95% CI:1.349,4.880).

## Discussion

In this study, the incidence of CHE post-pandemic among households in Negeri Sembilan was 16.8%, markedly higher than the national average which was approximately only 2.8% reported in 2016 (11). The escalation of CHE seemed to have intensified during the COVID-19 pandemic, as it has been well documented in two separate studies that reported varying incidence of CHE. A study that focussed on preventive measures recorded CHE at 8% (11), meanwhile a study focussed on community in Sarawak recorded much higher CHE at 20.2% reflected acute phase of pandemic (12,20). This wide gap figures between both studies may be largely influenced by differences in sample size and study context. The preventive study involved a small sample, while the Sarawak study included a much broader sample. Moreover, Sarawak, being a predominantly rural state, faces greater healthcare accessibility challenges and higher OOPHE. Although the CHE post-pandemic reported in this study was marginally lower than during the pandemic, both periods showed significant increased compared to the pre-pandemic phase. Reinforcing these concerns, a previous study indicates that Malaysia’s per capita health spending is expected to grow by 8.3% annually from 2023 to 2028 further signalling the financial burden (21).

Similar patterns of CHE from varies countries have been seen where there was an increased in CHE from pre-pandemic to pandemic phase, depending on their health system structure, financial protection mechanisms, and economic resilience during the pandemic. For instance, previous recorded that in Mexico CHE was 3.3% increased to 5.6% in 2020, meanwhile in Peru CHE recorded at 8.7% at 2019 to 9.6% to 2020 (22). In contrast with a study conducted Uganda, based on evidence from the 2019/2020 Uganda National Household Survey indicated a reduction in CHE, whereby CHE decreasing from 38.07% in the pre-pandemic to 37.28% during the pandemic (10). Nonetheless, this decline did not indicate enhanced financial protection, instead it was likely influenced by the avoidance of healthcare, as numerous households postponed treatment, evaded health facilities, or opted for home-based care due to financial limitations.

On the other hand, during the pandemic, most of the populations shift from public to private healthcare services as most public healthcare were overwhelmed, temporarily inaccessible during lockdowns and prioritized COVID-19 patients (22). The population consequently sought healthcare from private facilities that operate on a fee-for-service basis, resulting in increased dependence on OOPHE and greater financial burden. Furthermore, to mitigate the transmission of the pandemic, numerous individuals utilised non-pharmaceutical interventions, including face masks, hand sanitisers, and various protective items. Adapting to the new norm became essential during the peak of the pandemic and continues to be relevant in the post-pandemic context. However, the majority of these items lacked government subsidies, leading to increase OOPHE for households (20,23). A study conducted in Indonesia found out that average of 7.25% of household income lost only for preventive measures during the pandemic heighten the financial burden (23).

To the best of our knowledge, no published studies to date have reported on post-pandemic CHE in Malaysia. Notably, our findings revealed a marked increase in CHE during the post-pandemic period compared to pre-pandemic levels. The high incidence of CHE in the post-pandemic period can be partly attributed to ongoing medical cost inflation. According to Bank Negara Malaysia, in 2023, the country recorded a medical cost inflation rate of 12.6%, more than double the global average of 5.6% (24). The increased of medical inflation rate has contributes to the high CHE and continues in post-pandemic period (21). A substantial proportion of the population is still struggling to achieve socio-economic and good health, i.e., the low-income groups. The low-income group in Malaysia might bear a higher burden of out-of-pocket health expenditure (OOPHE) and health challenges, mainly related to noncommunicable diseases (NCD). The costs are sometimes high enough so that households cannot recuperate them from existing resources and lead them to financial problems. This study analysed the factors associated with OOPHE among B40 group in Malaysia using nationwide data from the 2015 National Health and Morbidity Survey (NHMS). Ordinary Least Square (OLS) regression analysis is used to study the factors associated with OOPHE. This study suggests that the gender (female) of household, income, T20 income group, tertiary education, unpaid worker, retire, and NCD raises the likelihood of incurring OOPHE. The result for a private employee, self-employed, and insurance status is spending less on OOPHE. Therefore, the results show a clear need for government action to enhanced primary healthcare programs and a focus on achieving Universal Health Coverage. This situation can protect the household from OOPHE financial burden and NCD, exceptionally low-income earners (25). Moreover, recent study identified a rise in healthcare utilization, with outpatient service used potentially surpassed pre-pandemic levels across both urban and rural settings in Malaysia. Although the pandemic has transitioned into an endemic phase, the financial burden associated with healthcare remains a major concern for many households (21).

Given the rising incidence of CHE post-pandemic, identifying its underlying determinants is essential to understanding the financial burden. Although high OOPHE is associated with catastrophic expenditure and impoverishment effect, it was important to note that not all high OOPHE resulted in CHE (11). For instance, findings from this study discovered that small household size were three times more likely at risk of CHE, even though the median of OOPHE was higher in larger family size. This outcome was similar to a pre-pandemic study (11). In our study, we found that smaller household was primarily composed of the elderly, who tended to utilize healthcare services more frequently due to higher healthcare demands, especially in the presence of chronic diseases. Several studies reported that households with the presence of elderly was a significant determinants of CHE (26–29). A study in Vietnam reported that most reported diseases among elderly consist of hypertension, arthritis, and musculoskeletal diseases with incidence of CHE at 49.5%, 20.6%, and 13.7% respectively borne notably higher OOPHE (29). Furthermore, with fewer income-contributing members, these households were more vulnerable to financial strain from healthcare spending (28).

This study found that heads of households with higher education had nearly three times the odds of experiencing CHE, a finding that contrasts with several previous studies (25–27). Typically, individuals with higher education are presumed to have greater health literacy and more likely to allocate resources on health due to heightened awareness (11,27). Consistent with previous findings, individuals with higher educational attainment in Malaysia demonstrate better health literacy and are more proactive in seeking quality healthcare services, reflecting improved health-related knowledge and attitudes (30,31). Nevertheless, higher education is not necessarily associated with higher income, as our study found that many of those with higher education did not belong to higher income category.

The study also indicated that household head in the non-working/retired category as significant determinants of CHE, that consistent with previous research (27). This was likely due to unstable and insufficient income to afford for healthcare. Further investigation identified that households head in this category comprised mainly of the elderly and those from the lower-income group, which heightened financial burden. Although Malaysia has dual-tier healthcare system, private facilities are often preferred by many Malaysians across various socioeconomic status, due to perceived higher quality, faster access, and more immediate service delivery (32). The findings from this study underscore the persistent financial vulnerability faced by Malaysian households in the post-pandemic era, particularly in Negeri Sembilan. The elevated incidence of CHE, could be explained with inflation in medical costs, increased healthcare utilisation, and socioeconomic disparities, reflects the multifaceted nature of healthcare-related financial hardship (21,24).

### Study limitations

This study is subject to several limitations. Firstly, the use of convenience sampling may affect the generalizability of the results and introduce potential selection bias. To mitigate this, the sampling procedure was proportionally guided by the population composition of each district in Negeri Sembilan. Secondly, the study is susceptible to recall bias, as participants were required to recollect household expenditures from the previous month, that may affect the accuracy of the data. Furthermore, the possibility of disclosure bias cannot be ruled out, as respondents may have intentionally misreported or withheld financial information due to privacy concerns or discomfort in disclosing sensitive household financial matters.

## Conclusion

The post-pandemic CHE incidence among households in Negeri Sembilan, Malaysia was at 16.8%, much higher than the pre-pandemic period. Lower household size, district’s location, higher educational level and lack of occupation were significant determinants of CHE. Findings from this study warrant the need for regular national monitoring of OOPHE and CHE through serial surveys. The identification of specific determinants highlights the need for targeted financial protection strategies. Strengthening public healthcare subsidies, expanding insurance coverage, and addressing structural inequalities will be essential to reduce the burden of CHE and promote greater equity in healthcare access in the post-pandemic recovery phase.

## Data Availability

Data cannot be shared publicly because it is the property of Universiti Sains Islam Malaysia and the Ministry of Higher Education. Data are available from the USIM Research and Innovation Management Centre (contact via https://pppi.usim.edu.my/) for researchers who meet the criteria for access to confidential data. The data underlying the results presented in the study are available from the USIM Research and Innovation Management Centre.

## Acknowledgement

This work was supported by Fundamental Research Grant Scheme (FRGS), Ministry of Higher Education, Malaysia under project number **FRGS/2/2022/SKK04/USIM/03/1**.

## Author Contributions

**Research grant principal investigator:** Mohamed Fakhri Abu Baharin

**Conceptualization**: Mohamed Fakhri Abu Baharin

**Data collection:** Siti Zulaikha Mohd Zainudin, Sharifah Najwa Syed Mohamad

**Data analysis**: Siti Zulaikha Mohd Zainudin, Nizam Baharom

**Supervision**: Mohamed Fakhri Abu Baharin, Nizam Baharom, Sharifah Najwa Syed Mohamad

**Writing– original draft**: Siti Zulaikha Mohd Zainudin

**Writing– review & editing**: Mohamed Fakhri Abu Baharin, Nizam Baharom, Sharifah Najwa Syed Mohamad

